# Sociodemographic inequalities in cardiovascular risk factors among adolescents from indigenous areas of Chiapas, México

**DOI:** 10.1101/2020.11.12.20230110

**Authors:** Elena Flores-Guillén, Itandehui Castro-Quezada, César Antonio Irecta-Nájera, Pilar E. Núñez-Ortega, Miguel Cruz, Roberto Solís-Hernández, Rosario García-Miranda, Paola Cruz-Cruz, Héctor Ochoa-Díaz-López

## Abstract

**Objectives:** The objective of this study was to determine the prevalence of cardiovascular risk factors among different sociodemographic and geographic areas of adolescents from indigenous areas of Chiapas, México.

**Design:** **A c**ross-sectional study.

**Setting:** **C**ommunities in the Totzil - Tseltal and Selva region of Chiapas, Mexico, were studied. Urban and rural areas of high marginalization according to the Human Development Index.

**Participants:** 253 adolescents were studied, of which 48.2% were girls and 51.8% were boys.

**Primary and secondary outcome measures:** a descriptive analysis of the quantitative variables was performed through central tendency and dispersion measures. Prevalence of cardiovascular risk factors and 95% confidence intervals (95% CI), stratified by sex, geographic area (rural/urban), schooling and ethnicity of mothers were estimated.

**Results:** the predominant risk factor in the study population was low HDL-c (51%). Higher prevalences of abdominal obesity and high triglycerides in girls were found and abnormal diastolic blood pressure in boys was identified. In urban areas were found greater prevalences of overweight/obesity and of insulin resistance while abnormal blood pressure levels were more prevalent in rural areas. Differences were found in the educational levels and ethnicity of the adolescents’ mothers. Prevalence of metabolic syndrome was 10% according to NCEP-ATPIII.

**Conclusions:** In this study, sociodemographic and geographical disparities were found in cardiovascular risk factors. Prevalence of risk factors was high, affecting mostly girls and urban population. Thus, there is a great need to promote healthy lifestyles and health, social and economic interventions to prevent chronic diseases in adulthood.

**Strengths and limitations of this study:**

- The value of this study lies on documenting sociodemographic inequalities in cardiovascular risk factors among population groups in marginalized Mayan communities of Chiapas, scarcely studied before.
- The sample included subjects from urban and rural areas.
- This was a cross-sectional study and causality cannot be inferred from our results.

| SUMMARY BOX |  |
| --- | --- |
| “What is already known on this subject?” | “What does this study add?” |
| Previous studies have shown that obesity and certain dyslipidemias are highly prevalent among adolescents from central, western and northern Mexico, but very little is known about southern Mexico. | Our study adds to the evidence suggesting that sociodemographic disparities are found in cardiovascular risk factors prevalence in adolescents from highly marginalized areas of Chiapas, Mexico |
| Sociodemographic inequalities affect cardiovascular risk. | Girls had higher prevalence of abdominal obesity, borderline total cholesterol and high triglycerides. Boys presented greater DBP levels. In urban areas, prevalences of overweight/obesity and of insulin resistance were higher than rural areas. Abnormal blood pressure levels were more prevalent in rural areas than in the urban ones. Differences were also found according to mother's schooling and ethnicity. |
| Evidence has shown that Metabolic syndrome prevalence varies within groups and geographical regions. | In our sample, the prevalence of metabolic syndrome was 10% according to NCEP-ATPIII. This figure was slightly higher in girls, in adolescents from urban areas and whose mothers speak an indigenous language. The mean carbohydrate intake in rural areas was 356 grams per day, while in urban areas was 322 grams per day. These figures are higher than the average for rural Mexican adolescents. |

## Background

In recent decades, there has been a growing worldwide interest in the study and reduction of inequalities in health. According to a recent study, in Mexico, some diseases were more frequent among people from low socioeconomic status such as: malnutrition, tuberculosis, (1) infant mortality, congenital infections, anemia, some malignancies, metabolic syndrome and diabetes, among others. (2) In Mexico, heart diseases are also the main cause of general mortality, representing an important public health problem. There is evidence of the association of different risk factors with these diseases. Among the main ones: abnormal lipid metabolism during childhood,(3) which is the leading cause of the formation of atheromatous plaques during adulthood.(4) In addition, alterations in levels of serum glucose, insulin, C-Reactive Protein (CRP) (5), blood pressure and waist circumference measurements are also associated with an increased risk of cardiovascular diseases. (6)

In Latin America, obesity and dyslipidemias are highly prevalent.(7) A study with 180 adolescents from Mexico City revealed that 61% had hypertriglyceridemia and half of them h low concentrations of high density lipoprotein cholesterol (HDL-c)(8). In Morelos, a study with 869 adolescents found that 7% had Metabolic Syndrome (MetS), 68% hypertriglyceridemia, 17% low HDL-c and 15% hyperglycemia. (9) In Tuxtla Gutiérrez, Chiapas, the prevalence of MetS in adolescents was 16%.(10) So far, there is no evidence in literature of the prevalence of cardiovascular risk factors in different sociodemographic groups of adolescents from Chiapas such as: sex, geographic area, ethnicity and education. Thus, the aim of this study was to present and analyze the distribution of cardiovascular risk factors among these populations from indigenous areas of Chiapas.

## METHODS

### Study design

A cross-sectional survey was carried out in a sample of adolescents obtained from a birth cohort study in three public hospitals in Chiapas in 2003. Sampling of the adolescents was undertook in two stages. In the first stage, the communities of the adolescents were clustered according to their size and geographical area; second stage, a systematic sampling of adolescents was carried out with a randomized start-up. (11)

### Study population

303 adolescents from 14 municipalities belonging to *Tzotzil-Tzeltal* and Selva regions of Chiapas were studied. (11) 50 participants without biochemical data were excluded, obtaining a final sample of 253 adolescents.

### Survey information

Interviews were conducted to adolescents and their mothers or caregivers at their homes. We applied a structured questionnaire with the following sections: sociodemographic data, non-pathological personal history, family medical history, anthropometric and clinical measures were taken. The sociodemographic data included: sex, age, geographic area, ethnicity and years of schooling of adolescents and their mothers, household goods and type of cooking fuel.

Family medical history included first and second degree of consanguineous relatives’ diseases: obesity, diabetes, high blood pressure and cardiovascular disease and non-pathological personal history: smoking and alcohol consumption.

Anthropometric and clinical assessment included: weight, height, waist circumference and blood pressure readings. Blood samples were taken from the antecubital vein for biochemical analysis.

### Data collection procedures

Data collection was carried out by a multidisciplinary team. (11, 19) Weight (kg) was measured by electronic scales (Model UM081, Tanita Corporation, accuracy ± 100 g, Tokyo, Japan). Height (m) was measured using stadiometers (SECA, accuracy ± 1 mm, Berlin, Germany). The body mass index (BMI) was calculated by dividing weight (kg) by the height squared (m). The BMI z-score was calculated with Anthro plus V 1.0.4 software. Nutritional status was classified: overweight/obesity [BMI z-score ≥1 standard deviation (SD)], normal weight (−1.9 to 1 SD) and thinness (≤-2 SD). Waist circumference was measured by anthropometric tapes (SECA, precision ± 1 mm, Berlin, Germany). The cut-off point for abdominal obesity was ≥80 cm in girls and ≥90 in boys, according to the International Diabetes Federation 2005 (IDF) (12)

Blood pressure was measured twice, using a digital monitor (Model ·CH-453, Citizen, Japan) while the adolescent were seated and after a 5-minute rest. Systolic Blood Pressure (SBP) or Diastolic Blood Pressure (DBP) was classified as normal (<90th percentile), normal-high (≥90 to <95th percentile), and high (≥95th percentile), according to the US Health National Institutes’ percentile tables for age and sex (13).

### Biochemical measurements

Blood samples were taken within 12 hours fasting from the antecubital fossa by venipuncture. Serum was obtained by centrifugation at 6000 rpm x 10 minutes. Determinations of serum glucose, triglycerides, total cholesterol, HDL-c and low-density lipoprotein cholesterol (LDL-c) were performed by photometric enzymatic methods (Diasys, Diagnostic System, Holzheim, Germany), in an automated analyzer (Vitalab Selectra E, Vitalab Scientific, Île-de-France, France). The quantification of insulin serum levels was performed using the immunoenzymatic method by magnetic separation (Adaltis Diagnostics, Milano, Italy) in the Eclectic analyzer (Adaltis Diagnostics, Milano, Italy) and the quantification of High Sensitive C-Reactive Protein (HS-CRP), was performed by nephelometry (Genius, Chenzhen, China) and brand reagents (Diasys, Holzheim, Germany).

### Diagnostic criteria

#### Metabolic alterations

Glucose levels were classified according to IDF (normal blood glucose <100 mg/dL, abnormal blood glucose 100-125 mg/dL).(12) For serum lipid levels, the cut-off points of National Cholesterol Education Program (NECP) were used: triglycerides (acceptable <90 mg/dL, borderline 90-129 mg/dL and high ≥130 mg/dL), total cholesterol (acceptable <170 mg/dL, borderline 170-199 mg/dL, high ≥200 mg/dL), LDL-c (acceptable <110 mg/dL, borderline 110-129 mg/dL, high ≥130 mg/dL) and c-HDL (acceptable >45 mg/dL, borderline 40-45 mg/dL, low <40 mg/dL) (14).

#### Insulin Resistance

The homeostatic model to assess insulin resistance (HOMA-IR) was calculated by multiplying fasting blood glucose (mg/dL) by fasting insulin (µU / mL) divided by 22.5. The cut-off point used to determine insulin resistance was 2.97.(15)

To define the degree of cardiovascular disease risk by HS-CRP, the cut-off points of the Center for Disease Control (CDC) and American Heart Association (AHA) were used: normal HS-CRP <1 mg/dL, moderate risk 1–3 mg/dL and high risk >3 mg/dL.

#### Metabolic Syndrome (MetS)

The diagnosis of MetS was analyzed under the criteria of IDF, Latin American Diabetes Association (ALAD)(16) and NCEP-Adult Treatment Panel III (NCEP-ATPIII). According to IDF, MetS is considered when there is abdominal obesity (≥90th percentile) and two or more of the following factors: triglycerides ≥150 mg/dL, HDL-c <40 mg/dL, SBP ≥130 or DBP ≥85 mmHg and glucose ≥100 mg/dL or previous diagnose of type 2 diabetes. (12)

The ALAD criteria are similar to those of IDF except for abdominal obesity, where different cut-off points were established according to sex and age. (16)

According to NCEP-ATPIII, three or more of the following factors must be included in the diagnosis of MetS: abdominal obesity (waist circumference ≥90th percentile both sexes), triglycerides ≥110 mg/dL, HDL-c <40 mg/dL, SBP or DBP> 90th percentile and glucose ≥110 mg/dL (17).

### Statistical analysis

A descriptive analysis of the quantitative variables was performed through central tendency and dispersion measures. Prevalence of cardiovascular risk factors and 95% confidence intervals (95% CI), stratified by sex, geographic area (rural/urban), schooling and ethnicity of mothers were estimated. Difference between distributions was obtained by Mann-Whitney U tests or *t*-tests for independent samples, while in the qualitative variables proportions were compared with Bonferroni correction. All analyses were performed in SPSS (version 23, 2018, SPSS Inc.).

## RESULTS

Adolescents’ sociodemographic characteristics according to sex are presented in Table 1. A total of 253 adolescents, 52% of the participants were male and 75% lived in urban areas. The average age was 14 years old and their average years of schooling, 6.5 years. 46% of adolescents’ mother spoke an indigenous language and 37.4% had <6 years of schooling. 30% of the adolescents’ father had less than 6 years of schooling and 47.2% spoke an indigenous language. It was identified that 4% of households did not have piped water, 31% cooked with charcoal/firewood, 36% did not have refrigerator, 12% without a television, 22% did not have mobile phone and 81% did not have computer. The population studied is mostly indigenous, with low schooling and living in precarious conditions, low availability of public services and poor housing conditions. Differences were found, although not statistically significant, between sexes by stratum and ethnicity.

**Table 1.** Sociodemographic characteristics of the sample of adolescents by sex^1^.

| Sociodemographic characteristics | Girls |  | Boys |  | Total |  | p-value |
| --- | --- | --- | --- | --- | --- | --- | --- |
|  | n | Mean ± SD or % | n | Mean ± SD or % | n | Mean ± SD o % |  |
| Age (years) | 122 | 14.1 ± 0.2 | 131 | 14.16 ± 0.3 | 253 | 14.15 ± 0.3 | 0.894<sup>3</sup> |
| Geographic area of residence |  |  |  |  |  |  |  |
| Urban | 95 | 77.9 | 94 | 71.8 | 189 | 74.7 | 0.311<sup>2</sup> |
| Rural | 27 | 22.1 | 37 | 28.2 | 64 | 25.3 |  |
| Adolescent´s schooling (years) | 122 | 7.7 ± 1.1 | 131 | 7.7 ± 1.0 | 253 | 7.7 ± 1.0 | 0.559<sup>4</sup> |
| Mother's schooling (years) | 118 | 6.8 ± 4.8 | 125 | 6.3 ± 4.4 | 243 | 6.5 ± 4.5 | 0.609<sup>4</sup> |
| 0 - 5 | 44 | 37.3 | 47 | 37.6 | 91 | 37.4 | 0.960<sup>5</sup> |
| 6 – 9 | 54 | 45.8 | 57 | 45.6 | 111 | 45.7 | 0.980<sup>5</sup> |
| >10 | 20 | 16.9 | 21 | 16.8 | 41 | 16.9 | 0.975<sup>5</sup> |
| Father´s schooling (years) | 105 | 7.9 ± 4.9 | 116 | 7.4 ± 4.3 | 221 | 7.6 ± 4.5 | 0.799<sup>4</sup> |
| 0 - 5 | 32 | 30.5 | 34 | 29.3 | 66 | 29.9 | 0.744<sup>5</sup> |
| 6 – 9 | 44 | 41.9 | 56 | 48.3 | 100 | 45.2 | 0.360<sup>5</sup> |
| >10 | 29 | 27.6 | 26 | 22.4 | 55 | 24.9 | 0.478<sup>5</sup> |
| Mother´s language (ethnicity) |  |  |  |  |  |  |  |
| No | 69 | 58.5 | 62 | 49.6 | 131 | 53.9 | 0.165<sup>2</sup> |
| Yes | 49 | 41.5 | 63 | 50.4 | 112 | 46.1 |  |
| Father´s language (ethnicity) |  |  |  |  |  |  |  |
| No | 55 | 55.0 | 58 | 50.9 | 113 | 52.8 | 0.584<sup>2</sup> |
| Yes | 45 | 45.0 | 56 | 49.1 | 101 | 47.2 |  |
| Household conditions |  |  |  |  |  |  |  |
| Television |  |  |  |  |  |  |  |
| No | 15 | 12.4 | 17 | 13.1 | 32 | 12.7 | 0.872<sup>2</sup> |
| Yes | 106 | 87.6 | 113 | 86.9 | 219 | 87.3 |  |
| Refrigerator |  |  |  |  |  |  |  |
| No | 36 | 29.8 | 54 | 41.5 | 90 | 35.9 | 0.065<sup>2</sup> |
| Yes | 85 | 70.2 | 76 | 58.5 | 161 | 64.1 |  |
| Microwave oven |  |  |  |  |  |  |  |
| No | 85 | 70.2 | 96 | 73.8 | 181 | 72.1 | 0.574<sup>2</sup> |
| Yes | 36 | 29.8 | 34 | 26.2 | 70 | 27.9 |  |
| Cell phone (head of household) |  |  |  |  |  |  |  |
| No | 25 | 20.7 | 30 | 23.1 | 196 | 21.9 | 0.651<sup>2</sup> |
| Yes | 96 | 79.3 | 100 | 76.9 | 55 | 78.1 |  |
| Computer |  |  |  |  |  |  |  |
| No | 96 | 79.3 | 106 | 81.5 | 202 | 80.5 | 0.750<sup>2</sup> |
| Yes | 25 | 20.7 | 24 | 18.5 | 49 | 19.5 |  |
| Cooking fuel |  |  |  |  |  |  |  |
| Gas | 42 | 34.7 | 30 | 23.1 | 72 | 28.7 | 0.042<sup>5</sup> |
| Firewood/coal | 29 | 24.0 | 51 | 39.2 | 80 | 31.9 | 0.010<sup>5</sup> |
| Both | 50 | 41.3 | 49 | 37.7 | 99 | 39.4 | 0.557<sup>5</sup> |

<sup>1</sup> Data are presented in means ± SD for quantitative variables and percentages for qualitative variables. <sup>2</sup> Fisher's exact test. <sup>3</sup> t-test. <sup>4</sup> Mann–Whitney U test. <sup>5</sup> Comparison of proportions test.

Table 2 describes the family medical history and non-pathological background of adolescents. Almost half of the adolescents had first or second degree relatives with chronic diseases. The most reported diseases were type 2 diabetes, hypertension and cardiovascular diseases. 32% of adolescents had an incomplete Mexican vaccination schedule and 6% were not breastfed during their first six months of life. 20% of adolescents consumed alcoholic beverages occasionally, and 16% had smoked. When analyzed by sex, boys showed a lower proportion than girls in the complete vaccination schedule and alcoholic beverages consumption.

**Table 2.** Family medical history and non-pathological personal history of adolescents^1^.

|  | Girls<br>(n=122) |  |  |  | Boys<br>(n=131) |  |  |  | Total<br>(n=253) |  |  |  |
| --- | --- | --- | --- | --- | --- | --- | --- | --- | --- | --- | --- | --- |
|  | n | % | CI 95% |  | n | % | CI 95% |  | n | % | CI 95% |  |
| **Family medical history<sup>1</sup>** |  |  |  |  |  |  |  |  |  |  |  |  |
| Obesity | 27 | 22.3 | 15.6 | 30.3 | 28 | 21.5 | 15.1 | 29.2 | 55 | 21.9 | 17.1 | 27.3 |
| Hypertension | 55 | 45.5 | 36.8 | 54.3 | 57 | 43.8 | 35.5 | 52.4 | 112 | 44.6 | 38.6 | 50.8 |
| Diabetes | 55 | 45.5 | 36.8 | 54.3 | 57 | 43.8 | 35.5 | 52.4 | 112 | 44.6 | 38.6 | 50.8 |
| Cardiovascular diseases | 54 | 44.6 | 36.0 | 53.5 | 57 | 43.8 | 35.5 | 52.4 | 111 | 44.2 | 38.2 | 50.4 |
| **Non-pathological personal history** |  |  |  |  |  |  |  |  |  |  |  |  |
| Complete vaccination schedule | 82 | 73.2 | 64.5 | 80.7 | 75 | 63.6 | 54.6 | 71.8 | 157 | 68.3 | 62.0 | 74.0 |
| Exclusive breastfeeding<sup>2</sup> | 110 | 91.7 | 85.7 | 95.6 | 119 | 95.2 | 90.4 | 98.0 | 229 | 93.5 | 89.9 | 96.1 |
| Have tried smoking | 18 | 14.9 | 9.4 | 22.0 | 22 | 16.8 | 11.2 | 23.9 | 40 | 15.9 | 11.8 | 20.8 |
| Have consumed alcoholic beverage | 30 | 24.6 | 17.6 | 32.8 | 20 | 15.3 | 9.9 | 22.2 | 50 | 19.8 | 15.2 | 25.0 |

<sup>1</sup> Family members of first and second degree of consanguinity (Parents and grandparents). <sup>2</sup> During the first six months.

The prevalence of cardiovascular disease risk factors by sex is shown in Table 3. The most prevalent risk factor was low HDL-c (51%), followed by overweight/obesity (29%), hypertriglyceridemia (29%) and insulin resistance (24%). Low prevalence of high LDL-c (1%), abnormal fasting blood glucose (4%) and high total cholesterol (4%) was identified. Girls had higher prevalence of abdominal obesity, borderline total cholesterol and high triglycerides. Boys presented greater DBP levels.

**Table 3.** Prevalence of cardiovascular disease risk factors in adolescents according to sex.

| Cardiovascular disease risk factors | Girls |  |  | Boys |  |  | Total |  |  | *p-value*<sup>1</sup> |
| --- | --- | --- | --- | --- | --- | --- | --- | --- | --- | --- |
|  | n | % | 95% CI | n | % | 95% CI | n | % | 95% CI |  |
| Anthropometric measures |  |  |  |  |  |  |  |  |  |  |
| Overweight/obesity (BMI >1 z-score) | 42 | 34.4 | 26.4 43.1 | 31 | 23.7 | 17 31.5 | 73 | 28.9 | 23.5 34.7 | 0.059 |
| Abdominal obesity (≥80 cm in girls and ≥90 cm in boys) | 30 | 24.6 | 17.6 32.8 | 7 | 5.3 | 2.4 10.2 | 37 | 14.6 | 10.7 19.4 | <0.001 |
| Clinical measures |  |  |  |  |  |  |  |  |  |  |
| SBP normal-high (≥90 percentile y <95 percentile) | 23 | 18.9 | 12.7 26.5 | 37 | 28.2 | 21.1 36.4 | 60 | 23.7 | 18.8 29.2 | 0.079 |
| SBP high (≥95 percentile) | 15 | 12.3 | 7.4 19 | 25 | 19.1 | 13.1 26.4 | 40 | 15.8 | 11.7 20.7 | 0.139 |
| DBP normal-high (≥90 percentile y <95 percentile) | 29 | 23.8 | 16.9 31.9 | 41 | 31.3 | 23.8 39.6 | 70 | 27.7 | 22.4 33.4 | 0.181 |
| DBP high (≥95 percentile) | 6 | 4.9 | 2.1 9.9 | 17 | 13 | 8 19.5 | 23 | 9.1 | 6 13.1 | 0.026 |
| Biochemical measures |  |  |  |  |  |  |  |  |  |  |
| Fasting blood glucose (≥ 100 mg/dL) | 2 | 1.6 | 0.3 5.2 | 6 | 4.6 | 1.9 9.2 | 8 | 3.2 | 1.5 5.9 | 0.182 |
| Borderline triglycerides (>90 - 129 mg/dL) | 40 | 32.8 | 24.9 41.4 | 48 | 36.6 | 28.8 45.1 | 88 | 34.8 | 29.1 40.8 | 0.520 |
| High triglycerides (≥130 mg/dL) | 43 | 35.2 | 27.2 44 | 29 | 22.1 | 15.7 29.8 | 72 | 28.5 | 23.2 34.2 | 0.021 |
| Borderline total cholesterol (170 - 199 mg/dL) | 18 | 14.8 | 9.3 21.8 | 5 | 3.8 | 1.5 8.2 | 23 | 9.1 | 6 13.1 | 0.002 |
| High total cholesterol (≥ 200 mg/dL) | 7 | 5.7 | 2.6 10.9 | 3 | 2.3 | 0.6 6 | 10 | 4 | 2 6.9 | 0.160 |
| Borderline LDL-c (110 - 129 mg/dL) | 7 | 5.7 | 2.6 10.9 | 3 | 2.3 | 0.6 6 | 10 | 4 | 2 6.9 | 0.057 |
| High LDL-c (≥130 mg/dL) | 2 | 1.6 | 0.3 5.2 | 0 | 0 | * * | 2 | 0.8 | 0.2 2.5 | * |
| Borderline HDL-c (40 - 45 mg/dL) | 28 | 23 | 16.2 31 | 28 | 21.4 | 15 29 | 56 | 22.1 | 17.4 27.5 | 0.763 |
| Low HDL-c (<40 mg/dL) | 54 | 44.3 | 35.7 53.1 | 74 | 56.5 | 47.9 64.8 | 128 | 50.6 | 44.5 56.7 | 0.052 |
| HOMA-IR (≥ 2.97 units) | 34 | 28.3 | 20.9 36.8 | 26 | 19.8 | 13.7 27.3 | 60 | 23.9 | 18.9 29.5 | 0.115 |
| HS-CRP moderate risk (1 - 3 mg/dL) | 20 | 16.5 | 10.7 23.9 | 13 | 9.9 | 5.7 15.9 | 33 | 13.1 | 9.4 17.7 | 0.120 |
| HS-CRP high risk (>3 mg/dL) | 17 | 14 | 8.7 21.1 | 13 | 9.9 | 5.7 15.9 | 30 | 11.9 | 8.3 16.3 | 0.312 |

Abbreviations: BMI, body mass index; SBP, systolic blood pressure; DBP, diastolic blood pressure; HOMA-IR, homeostatic model to assess insulin resistance; HS-CRP, high sensitivity C-reactive protein. \* Not calculable, a column with a proportion equal to zero. <sup>1</sup> Comparison of proportions using Bonferroni correction.

The prevalence of cardiovascular disease risk factors according to geographic area of residence (urban/rural) are shown in Table 4. In urban areas, a higher prevalence of overweight/obesity (32.8%) and insulin resistance (27.5%) were observed. In rural areas the normal-high DBP prevalence (40.6%) was higher than in urban ones.

**Table 4.**
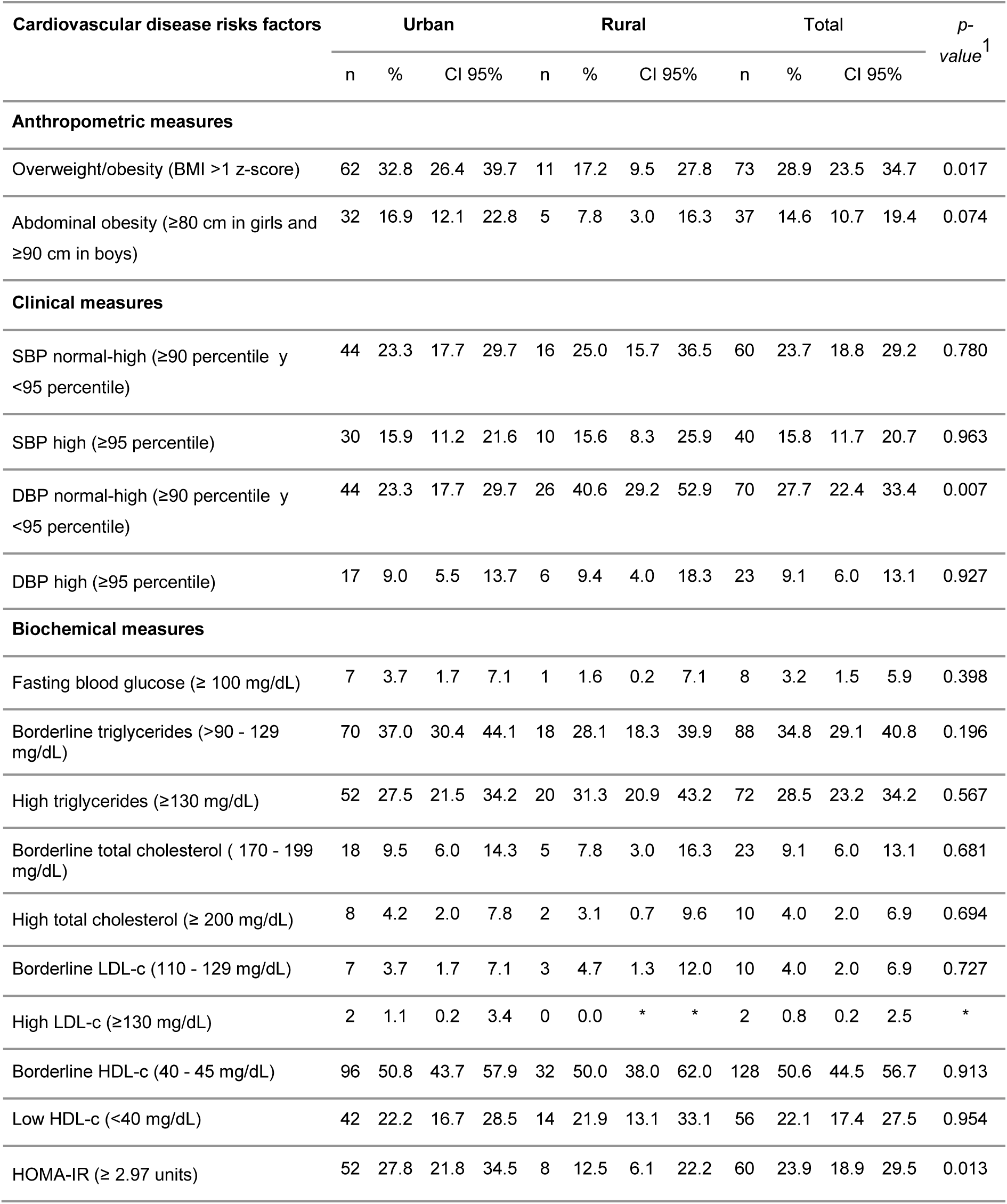
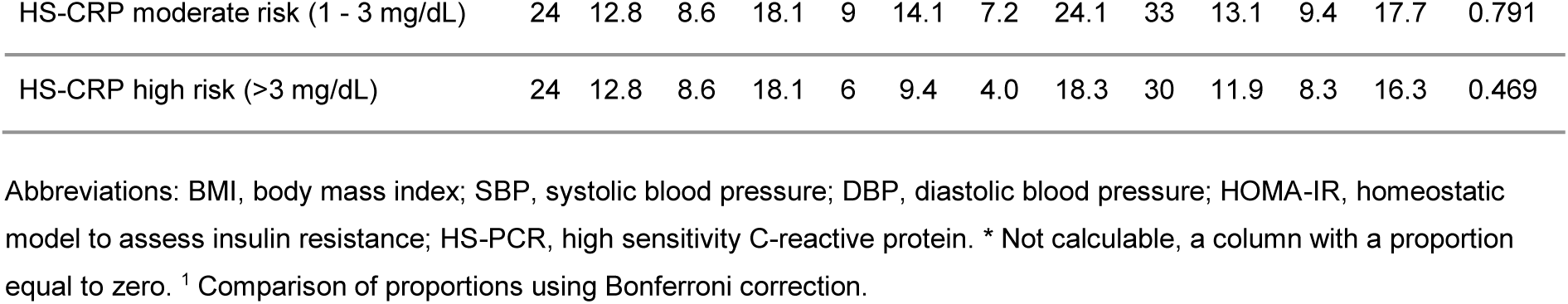
Prevalence of cardiovascular disease risk factors in adolescents according to geographic area of residence.

According to mother’s schooling, higher prevalence of overweight and obesity, abdominal obesity and serum lipid levels were identified in adolescents among mothers with schooling ≥7 years. In adolescents with mothers with less schooling, greater alterations in blood pressure, glucose, triglycerides and low HDL-c were observed.

Regarding mother’s ethnicity, there is a greater tendency to have risk factor alterations in adolescents whose mothers speak an indigenous language, except for obesity, triglycerides and borderline cholesterol, LDL-c, and insulin resistance. (Supplementary Tables 1 and 2).

The highest prevalence of MetS (10%) was according to NCEP-ATPIII, followed by IDF (3%) and ALAD (1%). Table 5 shows the MetS prevalence defined by NCEP-ATPIII criteria, according to different population groups. A higher prevalence of MetS was observed in girls, in urban areas and in adolescents with indigenous speaker mothers.

**Table 5.** Metabolic syndrome prevalence according to different sociodemographic groups and geographic areas of adolescents from Chiapas, Mexico.

| Variables |  | Metabolic syndrome |  | *p*-value<sup>1</sup> |
| --- | --- | --- | --- | --- |
|  |  | n | % |  |
| Sex | Girls | 13 | 10.7% | 0.690 |
|  | Boys | 12 | 9.2% |  |
| Geographic area | Urban | 21 | 11.1% | 0.337<sup>2</sup> |
|  | Rural | 4 | 6.3% |  |
| Mother's schooling (years) | 0 - 5 | 7 | 7.7% | 0.656 |
|  | 6 - 9 | 12 | 10.8% |  |
|  | >10 | 5 | 12.2% |  |
| Mother's language (ethnicity) | Spanish | 11 | 8.4% | 0.403 |
|  | Indigenous (Mayan) | 13 | 11.6% |  |
| Father's language (ethnicity) | Spanish | 11 | 9.7% | 0.781 |
|  | Indigenous (Mayan) | 11 | 10.9% |  |

<sup>1</sup> Chi squared test. <sup>2</sup> Fisher's exact test.

## Discussion

In this study, we found a high prevalence of cardiovascular risk factors such as overweight/obesity and abdominal obesity, diastolic blood pressure, cholesterol total borderline, HDL-c and insulin resistance, among adolescents from indigenous communities of two regions from Chiapas.

The prevalence of overweight/obesity in our study was 29%. This prevalence was higher in urban areas (32.8%). Similarly, the National Survey of Health and Nutrition of Mexico 2016 (ENSANUT) for adolescents (12 to 19 years) showed that overweight/obesity prevalence in urban areas was slightly higher (36.7%) than in rural areas (35%). Our data are similar to those presented in ENSANUT-2016. Over the last decades, obesogenic environment has been widely studied, especially in urban areas but there are scarce studies in rural areas, particularly among indigenous communities. Overweight/Obesity are the result of complex interactions between biological and behavioral individual factors and community environmental factors (socioeconomic, cultural practices, food availability and marketing, etcetera). In urban areas, Mexican population is mainly physically inactive (2) and often exposed to low-nutrient energy-dense foods and sugar-sweetened beverages.(18) Paradoxically, the double burden of malnutrition was present in our sample, as we already stated in a previous publication, a high proportion of the adolescents under study presented stunting in rural areas, rather than obesity.(11)

In our study, the general prevalence of abdominal obesity was 14.3%. We found differences by sex, girls with the highest prevalence (24.6%). In New Zealand, female adolescents have higher peripheral percentages of body fat (19). However, previous studies in Spain (20) and Brazil (21) presented a higher prevalence of abdominal obesity in boys, but the relationship between abdominal obesity and sex in adolescents is still controversial. The difference in body fat distribution between sexes may explain the higher percentage of body fat in girls, since there is a redistribution of fat from the extremities to the trunk. This distribution differs between sexes, because these changes are associated with the levels of estrogen and testosterone. (22) The disparities between sexes could be attributed to differences in sociodemographic, cultural and lifestyle factors such as physical activity, occupation and diet. Since 2012, prevalence of overweight in girls has risen from 23.7% to 26.4%, whereas in boys, no changes were observed.(23) In Chiapas, overweight/obesity prevalence in female adolescents has also increased from 30.7% in 2006 to 32.1% in 2012. (24)

Regarding blood pressure levels, in our study the proportion of boys with DBP ≥95 percentile was higher than girls. Previous research with adolescents in USA, also showed that after the onset of puberty, boys had a higher blood pressure than girls at same age (25). The mechanisms responsible for sex differences in blood pressure during adolescence are still unclear. There is evidence that androgens, such as testosterone, could play an important role in blood pressure regulation in sex-related differences,(26) since testosterone levels cause endothelial dysfunction (27). In rural areas, a higher prevalence of normal-high DBP was found in our study. This finding contrasts with other study conducted in Mexico City and rural areas of the State of Mexico (28) where higher blood pressure levels were found in urban areas. According to our results, some discretionary foods were more frequently consumed in rural areas, i.e. carbonated beverages (81.8% once a week or more in rural areas vs. 59.7% in urban areas) and chips (35.1% >3 times per week in rural areas vs. 25.7% in urban areas).

Our findings related to blood lipids, analysis stratified by sex showed that triglycerides were higher in girls than in boys. These differences by sex could be explained by their different hormonal activity during adolescence. (29) Girls of our study had higher values of waist circumference which might explain their higher values of triglycerides. The most prevalent risk factor found in rural areas was a high level of triglycerides, although this difference with urban areas was not statistically significant. This result is in accordance with those reported in a study among adolescents from Mexico City and a rural community in central Mexico, where 45% of the population are predominantly *Mazahuas and Otomíes* peoples. (28) Low-fat and high-carbohydrate diets increase triglyceride levels (30). In our study, the mean carbohydrate intake in rural areas was 356 grams per day, while in urban areas was 322 grams. These figures are higher than the average for Mexican rural adolescents, (285 grams per day in girls, 326 grams in boys). Furthermore, it has been reported that 69% of female adolescents and 61% of male in rural areas consume an excess of added sugars. (31)

We also found that the prevalence of borderline total cholesterol was higher in girls (14.8%) than in boys (3.8%) but we did not find differences by geographic area. It has been described that total cholesterol levels decline in males in early puberty but rise again as males approach adulthood. In contrast, total cholesterol levels increase throughout puberty in females. (32) However, we did not measure Tanner stages, therefore we could not differentiate sexual maturation, which might influence our results. There are differences in levels of low HDL-c in mothers <6 years of schooling compared to those with >7 years.

The prevalence of insulin resistance in our study was considerably high (24%). In addition, it was found that insulin resistance in urban areas was higher than in rural areas. This finding agrees with previous studies where the urban environment was an independent predictor of insulin resistance. (33) Furthermore, there is evidence that insulin resistance is associated with lipid alterations in early life and overweight/obesity. (34) Thus, our results might also be explained by the high prevalence of obesity and alterations in lipid metabolism among our sample. No differences were found between sexes, ethnic groups and mothers’ education levels.

According to the HS-CRP analysis, it was found that 25% of adolescents under study had moderate and high cardiovascular risk according to CDC and AHA, higher proportions of HS-CRP are observed in girls, urban areas and adolescents whose mothers have less schooling, although there were no statistically significant differences. Otherwise, a research conducted with 418 Mexican adolescents from Guadalajara showed no gender differences in cardiovascular risk by HS-CRP. (35)

Regarding the MetS in the study population, the results showed the highest prevalence (9.9%) when measured by NCEP-ATPIII criteria. Other studies conducted in Mexico have shown different prevalences: 8.8% nationwide, (36) 6.7% in Morelos (9) and 20% in Campeche, (37) which has the highest prevalence of diabetes and hypertension at national level.

Among the possible explanations for the situation found in our study might be the nutritional transition among the population of Chiapas (38). Dietary patterns have shifted from traditional plant based diet to a westernized pattern consisting of high fat and refined carbohydrate intake observed in the last decades. (39) ENSANUT 2016 showed that adolescents commonly consumed food groups not recommended for daily consumption: 84% habitually consumed sugary drinks, 59% snacks, sweets and desserts and 50% sweetened cereals. (23)

Sedentary behavior among adolescents has also been associated with insulin resistance, lipid alterations and higher inflammatory states. (40) The adolescents studied reported to pass 10 hours per day seated (9 hours for rural population) and only 1 hour 40 minutes per week doing exercise or sports, no differences were found according to sex (data shown in supplementary tables).

Socioeconomic factors and mechanisms such as: a different access to food, education, health services, information, income, occupation and empowerment might explain the disparities found between population groups and geographical areas. The value of this study lies on documenting sociodemographic inequalities in cardiovascular risk factors among population groups in marginalized Mayan communities of Chiapas, scarcely studied before. This could allow identifying vulnerable groups to develop future interventions to modify their cardiovascular risk profiles.

## Conclusions

In this study with adolescents from marginalized indigenous areas of Chiapas, sociodemographic and geographical disparities were found in cardiovascular risk factors. It is noteworthy that girls had higher risk factors prevalences. A differential pattern of risk factors was found in geographical strata, urban population being more vulnerable. Thus, there is a great need to promote healthy lifestyles and other health, social and economic interventions in these adolescent groups to prevent future chronic diseases in adulthood.

## Data Availability

Any data could be available from the corresponding author on a reasonable request from the Editorial Office of the Journal.

## Declarations

### Ethics approval and consent to participate

Prior to the interview, mothers or caregivers and adolescents signed an informed consent. This study was approved by the Research Ethics Committee of El Colegio de la Frontera Sur (CEI-O-076/16).

## Consent for publication

Not applicable.

## Availability of data and materials

The datasets generated and analyzed during the current study are not publicly available due to our Federal Laws about copyright from the research work’s results resulting from ongoing projects as a Mexican public research center. Although, any data could be available from the corresponding author on a reasonable request from the Editorial Office of the Journal.

## Competing interests

The authors declare that they have no competing interests.

## Funding

This study was supported by the National Council for Science and Technology of Mexico (CONACYT). EFG received a PhD scholarship from PRODEP and PNO received a PhD scholarship from CONACYT. IC-Q received a postdoctoral scholarship from CONACYT.

## Authors’ contributions

HODL conceived and directed the project. EFG, HODL and CAIN designed and planned the study. EFG and PNO supervised the data collection. RSH performed the statistical analysis. HODL, EFG, PNO and ICQ contributed to the analysis and interpretation of the results. EFG, ICQ and HODL wrote the manuscript with contributions from all authors. MC, RGM, PC revised the manuscript with important contribution of all co-authors. All the authors approved the final version.

## Acknowledgements

The authors want to thank the participants of the study and the contribution of social service students from Universidad de Ciencias y Artes de Chiapas, Universidad del Sureste and Instituto de Estudios Superiores de Chiapas.

## Notes

### Competing Interest Statement

The authors have declared no competing interest.

### Author Declarations

This study was approved by the Research Ethics Committee of El Colegio de la Frontera Sur (CEI-O-076/16).

